# Comparing Machine Learning Methods for Detecting Caregiver Burden in Transcribed Conversations with Clinicians

**DOI:** 10.64898/2026.10.30.26366925

**Authors:** Ari Z. Klein, Weiqing He, Hongzhuo Chen, Boning Tong, Emily Summerhayes, Li Shen, Nancy A. Hodgson

## Abstract

Dementia caregivers’ elevated burden is associated with earlier patient institutionalization and greater caregiver morbidity, yet no standardized, objective method exists for detecting burden during routine clinical encounters. We compared four machine learning and AI approaches for detecting caregiver burden in de-identified caregiver-clinician transcripts from the COPE in PACE study using Zarit burden scores. A naive baseline predicting burden in every transcript achieved precision/F1 = 0.40/0.57. LLM prompting achieved precision/F1 = 0.43/0.60. Weak supervision achieved precision/F1 = 0.48/0.58. The fully supervised model, trained on sentence-level labels with only 1.1% burden-positive sentences, achieved precision/F1=0.19/0.25; this result is not directly comparable because of different labels and units of analysis. LLM prompting modestly outperformed weak supervision and naive baseline without fine-tuning, while class imbalance limited fully supervised sentence-level classification. Larger annotated corpora and refined labeling strategies are needed for clinical deployment.

## Introduction

Dementia caregivers are essential individuals who aid persons living with dementia (PLWD) with social activity and healthcare coordination such as transportation, appointment scheduling, and medication management, among other things^1,2^. These informal or unpaid adult caregivers provide support to relatives, friends, partners, or other individuals with whom they have a close personal relationship^3^. Caregiver burden is associated with higher health care costs, including early institutionalization of the PLWD and higher rates of caregiver mortality^4,5,6^. Compared with non-dementia caregivers, dementia caregivers are significantly more stressed and suffer from greater burden^7,8^.

When caregivers accompany PLWD to healthcare appointments, clinically valuable and relevant information can be extracted from conversations between the caregiver and the clinician^9^. Because the caregiver is not the direct patient, this interaction is traditionally a clinician’s most important source of information about a caregiver’s wellbeing. To obtain valuable information and flag concerning content indicating a caregiver’s level of burden, clinicians require sufficient consultation time and strong clinical competencies^10^. However, no standardized, objective measure exists for detecting these changes during a clinical conversation, leaving considerable room for speculation and subjective bias^11^.

To address caregivers’ wellbeing during PLWD healthcare visits, objective markers of caregiver distress such as burden are necessary for improved identification and treatment. Given the current rise of artificial intelligence (AI) in healthcare, we evaluated technology-based methods in data science, including natural language processing (NLP) and machine learning (ML), for measuring clinical communications, making inferences about burden level, and guiding treatment and care plans^12^.

## Methods

### Data collection

This study is a secondary analysis of audio-recorded and de-identified conversations and clinical assessment data collected during a larger implementation study evaluating the translation of an evidence-based dementia program into a long-term care setting. The COPE in PACE study (NCT04165213) compared two training approaches for the Care of Older Persons in their Environment (COPE) program in improving interventionists’ uptake of COPE principles^13, 14, 15^. COPE was implemented in 10 Programs of All-Inclusive Care for the Elderly (PACE) across the United States. Through the PACE program, enrollees in the local community are guided by an interdisciplinary healthcare team and have access to most long-term care services covered by Medicaid and Medicare, including adult day care, home care, transportation, and healthcare services^6, 16^.

Parent-study participants included occupational therapists and registered nurses (n = 15) delivering the COPE program to dyads (n = 17) consisting of caregivers and PLWD. Among other outcome measures, caregivers completed the 12-item Zarit Burden Interview at baseline and follow-up to COPE.^17^ Throughout COPE delivery, clinicians delivered and recorded between 1 and 8 sessions with caregivers over the course of 3 to 8 weeks. Each session lasted one hour on average. In total, 126 COPE sessions were recorded, resulting in over 150 hours of conversation between COPE clinicians and caregivers.

To prepare the dataset for machine learning, a team of research assistants listened to recordings and flagged protected health information (PHI) (i.e., names, addresses, phone numbers). Audio files were de-identified by placing a beep tone over spoken instances of PHI before being automatically transcribed by secure speech-recognition software. Resulting transcripts were further de-identified by reviewing and removing any additional potentially identifying information.

The de-identified transcripts, annotation labels, and derived analysis datasets used in this study contain sensitive clinical information collected under an IRB-approved protocol and are not publicly available. De-identified data may be made available from the corresponding author upon reasonable request and execution of a data use agreement, subject to institutional and IRB approval.

### Classification

The dataset for this study consists of recorded, transcribed, and de-identified sequential discussions between dementia family caregivers and clinicians, together with Zarit Burden Interview scores completed by caregivers at baseline. We developed and evaluated four ML and AI approaches to classify caregiver speech signaling high versus low caregiver burden: two at the transcript level and two at the sentence/conversational turn level.

### Method 1: Weak supervision (transcript level)

At this point in the study, 98 of the 126 audio recordings had been transcribed. These 98 transcripts were split into three sets at the caregiver level: 45 transcripts for training (23 transcripts from 2 high-burden caregivers and 22 transcripts from 3 low-burden caregivers), 23 transcripts for validation (10 transcripts from 2 high-burden caregivers and 13 transcripts from 2 low-burden caregivers), and 30 transcripts as held-out test data (12 transcripts from 2 high-burden caregivers and 18 transcripts from 2 low-burden caregivers). In the first approach, a classifier was trained at the sentence level, where sentences were labeled using the caregiver’s overall Zarit burden score from the parent study; that is, every sentence from a given transcript inherited the caregiver’s baseline high- or low-burden label determined by the Zarit cutoff scores a priori^17^. A deep neural network classifier was trained by fine-tuning a pre-trained transformer model in the Hugging Face^19^ library (BERT-Base-Uncased)^20^ using 5 epochs, a batch size of 8, and a learning rate of 1e-5. The validation set was used to evaluate the model after each epoch, and the best-performing model was re-loaded to predict labels for the 45,245 sentences in the test set. A majority-based voting heuristic was used to aggregate the predicted labels for transcript-level evaluation.

### Method 2: LLM prompting (transcript level)

In this approach, we evaluated transcript-level LLM prompting using two large language model systems: Qwen3-30B-A3B-Instruct-2507,^21^ an instruction-tuned large language model deployed locally using the Hugging Face Transformers framework, and PennDnA, a Penn-managed GPT-based large language model system accessed through an API, in which study data are not transmitted to OpenAI-hosted services. The same prompting and evaluation procedure was applied to both systems. De-identified transcripts were formatted as speaker-attributed dialogue, with speakers identified as healthcare providers or caregivers of persons living with dementia. The prompt defined caregiver burden as the negative consequences of the time and effort spent providing care, including personal, role, financial, and social strain. The model was instructed to begin its response with “yes” and provide supporting quotations when burden was present or respond only with “no” when burden was not identified. Each transcript was analyzed independently without model training or fine-tuning. The initial yes/no response was treated as the transcript-level prediction and evaluated against the held-out set of 12 high-burden and 18 low-burden transcripts.

### Method 3: Full supervision (conversational turn level)

In the third approach, a gold-standard corpus was curated to train and evaluate the classifiers. Annotation guidelines were developed to help 3 of the annotators (HC, HR, and FY) distinguish between sentences that did and did not indicate high caregiver burden, based on the operational definition of caregiver burden as defined by the Zarit Burden Interview. For annotation, the annotators listened to the audio recordings while reading the transcripts. All 126 transcripts were manually annotated, regardless of the caregivers’ Zarit scores. After dually annotating 45 transcripts to establish an inter-annotator agreement (Cohen’s kappa) of 0.79, 22 the remaining 81 transcripts were each annotated by one annotator. Among the 191,508 total sentences, 2102 (1.1%) indicated high caregiver burden and 189,406 (98.9%) did not—a highly imbalanced dataset^23^. To capture additional context, all the sentences and annotated labels from a speaker’s conversational turn (i.e., before another speaker begins) were then merged into one unit of text. Among the 54,497 total conversational turns, 1078 (2%) indicated high caregiver burden and 53,419 (98%) did not. The same transcripts from the previous approaches were used in the training, validation, and test sets, and the remaining 28 transcripts were added to the training set. Again, a deep neural network classifier was trained and evaluated by fine-tuning the BERT-Base-Uncased^20^ pre-trained transformer model, using 5 epochs, a batch size of 8, and a learning rate of 1e-5. To address the class imbalance^23^, additional experiments were performed in which the (1) the minority class was oversampled, (2) the majority class was randomly under sampled, and (3) the scikit-learn Python library^23^ was used to automatically compute class weights, which assign more importance to, and a higher penalty for misclassifying, the minority class. In addition, two loss functions designed for long-tailed classification were evaluated: (4) Vector-Scaling (VS) loss^24^, which applies class-dependent multiplicative scaling and additive bias terms to the output logits before computing the loss, thereby adjusting the effective decision boundaries according to class imbalance, and (5) Label-Distribution-Aware Margin (LDAM) loss^25^, which enforces larger classification margins for minority classes by subtracting a class-specific margin from the true-class logit during training. For both loss functions, the governing hyperparameters (multiplicative and additive logits for VS loss; maximum margin and logit scale for LDAM loss) were tuned using Bayesian hyperparameter optimization with Optuna^26^.

### Method 4: LLM prompting (conversational turn level)

To evaluate LLM prompting at a finer level of granularity, we additionally applied the PennDnA system to the manually annotated conversational-turn data. Using the same general burden-classification prompting framework described above, we evaluated two input strategies: single-turn prompting and context-augmented turn-level prompting. In the single-turn approach, each annotated conversational turn was submitted to the model independently, without surrounding conversational context. In the context-augmented approach, each target turn was accompanied by up to four preceding conversational turns from the same conversation. The model was instructed to use the preceding turns only as contextual information and to determine whether the target turn itself indicated caregiver burden. In both strategies, the required output was a binary “yes” or “no” response, which was parsed as burden or no burden, respectively, and compared with the corresponding manually annotated turn-level reference label. Responses that could not be parsed as either “yes” or “no” were excluded from performance evaluation. However, no such responses occurred in the final evaluation.

### Evaluation metrics

Model performance was evaluated using precision, recall, and F1-score for the class that indicated high caregiver burden (“positive” class). They were quantified using standard binary classification metrics, computed over true positives (TP), false positives (FP), and false negatives (FN):

Precision = TP / (TP + FP)

Recall = TP / (TP +N)

F1 = 2 × (Precision × Recall) / (Precision + Recall)

Methods 1 and 2 were evaluated at the transcript level, using the caregivers’ Zarit scores as ground truth for whether the transcripts represented high burden or low burden. For Method 1, a transcript was considered to be a true positive if the caregiver’s Zarit score corresponded to high burden and the classifier predicted the majority of sentences as indicating high burden. For Method 2, a transcript was considered to be a true positive if the caregiver’s Zarit score corresponded to high burden and the LLM detected as least one sentence that indicated high burden. The performance of these methods was compared to a baseline classifier that would simply predict every transcript in the test set as high burden: precision of 0.40 (18 false positives), recall of 1.00 (no false negatives), and an F1-score of 0.57. In contrast, Method 3 and Method 4 were evaluated at the conversational turn level, using manually annotated labels as ground truth, rather than the caregivers’ Zarit scores. Because the unit of analysis and label source differ between these approaches, F1-scores are not directly comparable across Method 3 versus Methods 1–2, as noted in the Results and Discussion.

## Results

Table 1 and Figure 1 summarize model performance across the naive baseline and the three ML approaches.

**Table 1.** Performance of transcript-level machine learning and LLM prompting approaches for detecting high caregiver burden, using the caregivers’ Zarit scores as ground truth.

| **Model** | **Precision** | **Recall** | **F1-score** |
|---|---|---|---|
| Baseline | 0.40 | 1.00 | 0.57 |
| BERT-Base-Uncased | 0.48 | 0.77 | 0.58 |
| Llama-3.1-8B | 0.39 | 0.92 | 0.55 |
| Qwen3-30B | 0.40 | 1.00 | 0.57 |
| GPT-4.1-Mini | 0.46 | 0.83 | 0.59 |
| GPT-5-Mini | 0.43 | 1.00 | 0.60 |

**Figure 1.**
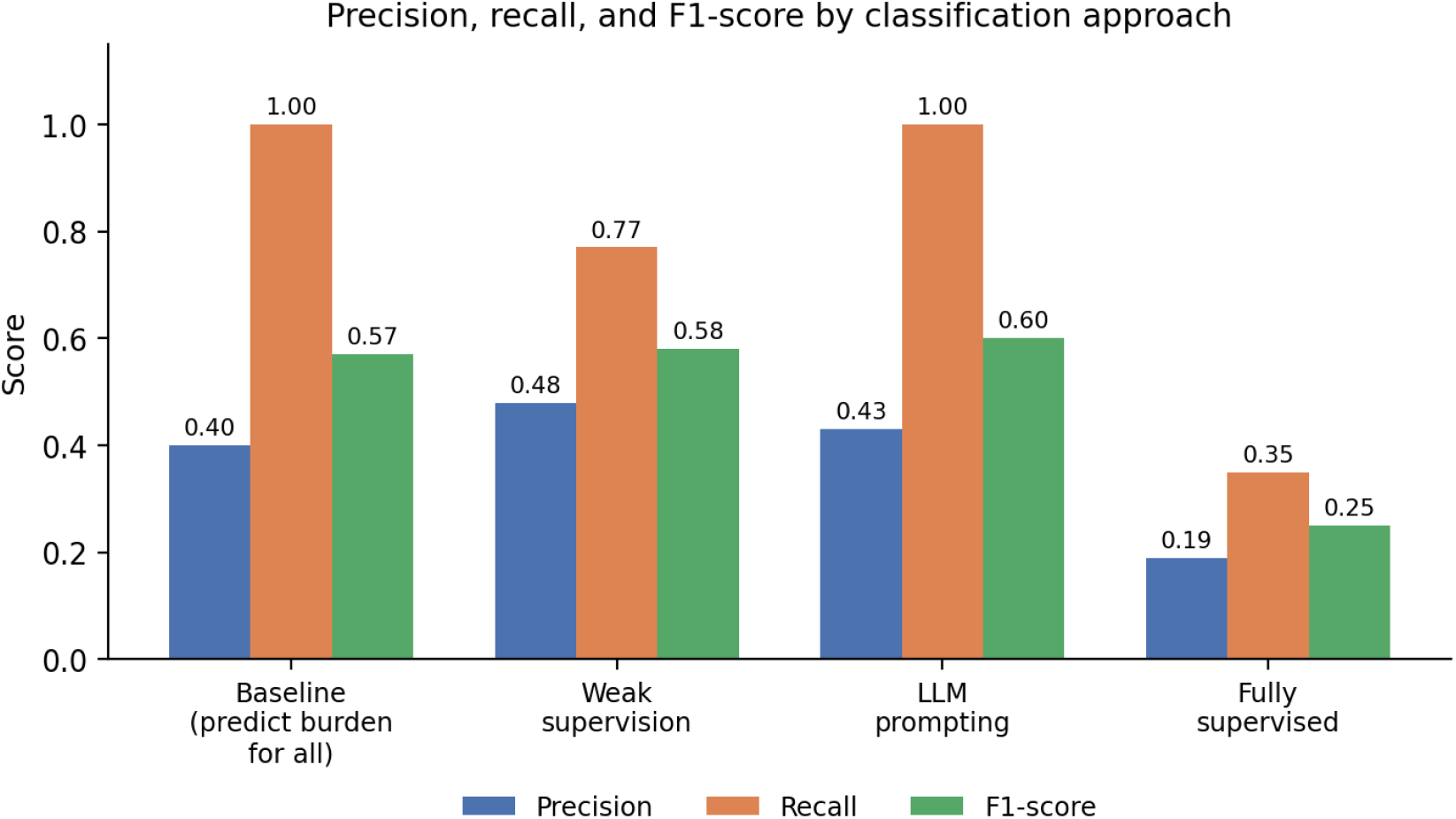
Precision, recall, and F1-score for the baseline and three burden-detection approaches.

Our LLM prompting approach achieved a recall of 1.00 and a precision of 0.43 (16 false positives), corresponding to an F1-score of 0.60 — the highest of the transcript-level approaches evaluated. Our weak supervision approach achieved higher precision (0.48) than both the baseline and LLM prompting, but this came at the cost of lower recall (0.77), yielding an F1-score of 0.58, which was only marginally higher than the baseline and lower than LLM prompting.

When we evaluated the fully supervised approach using manually annotated sentence-level labels rather than caregiver-level Zarit scores, we found that only 2,102 sentences (1.1%) of the 191,508 total sentences actually indicated burden. This sparse and imbalanced distribution presented substantial challenges for model training: the fully supervised model achieved an F1-score of 0.25 (precision = 0.19, recall = 0.35). This performance is not directly comparable to that of the first two approaches, because it used different labels (sentence-level human annotation rather than caregiver-level Zarit scores) and a different unit of analysis for evaluation (sentence vs. transcript). Table 2 summarizes the performance of detecting burden at the conversational turn level.

**Table 2.**
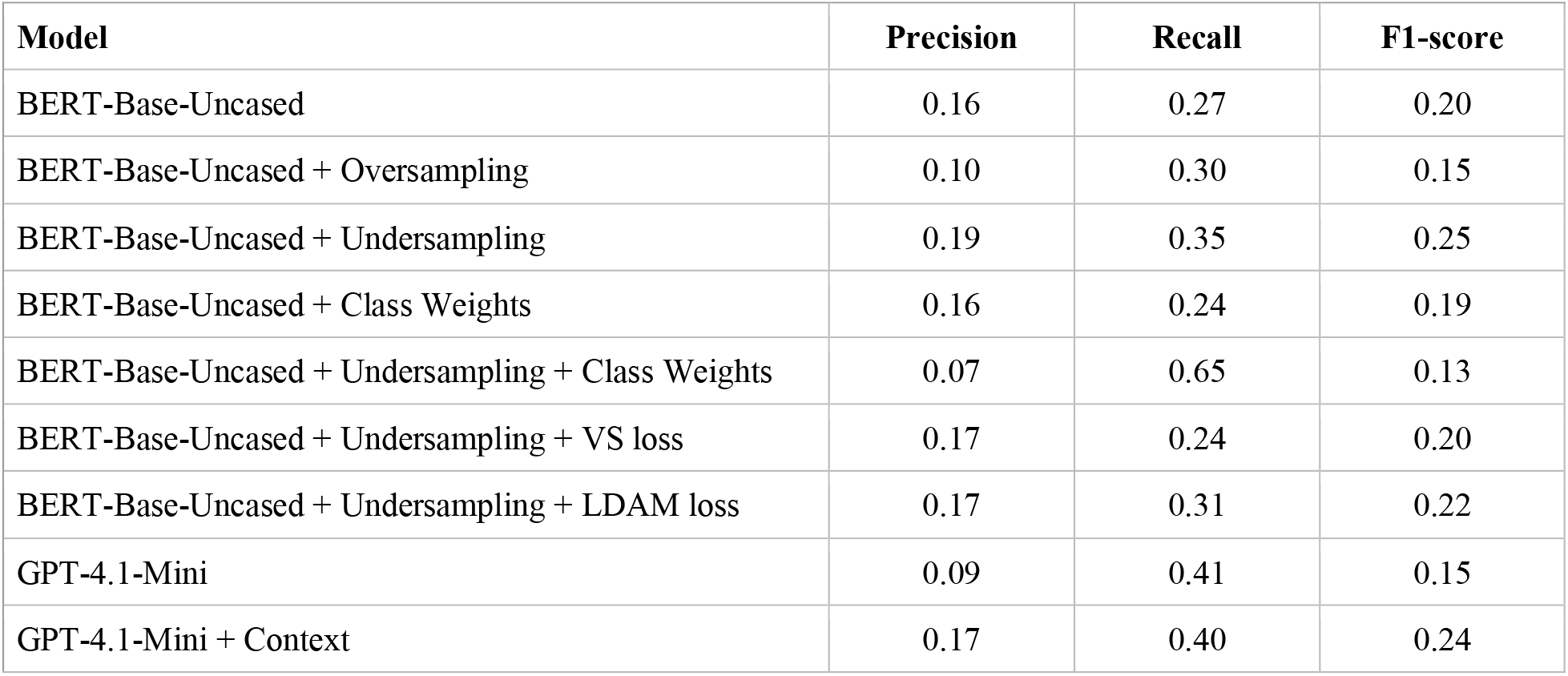
Performance of conversational turn-level machine learning and LLM prompting approaches for detecting high caregiver burden, using manually annotated labels as ground truth.

## Discussion

This study compared three ML approaches for detecting caregiver burden in transcribed clinical conversations, spanning a spectrum from weakly labeled to fully human-annotated supervision. Two findings stand out. First, LLM prompting, which required no model training or fine-tuning, only a structured description of the burden construct, achieved the highest F1-score among the transcript-level approaches, modestly exceeding both weak supervision and a naive baseline. This suggests that instruction-following LLMs may be able to encode clinically meaningful constructs, such as the Zarit Burden Interview’s definition of burden, directly from a prompt, offering a potentially low-cost complement to traditional supervised NLP pipelines in resource-constrained clinical NLP settings.

Second, the weak supervision approach illustrates a common trade-off in distantly supervised learning: because every conversational turn in a transcript inherited that caregiver’s single baseline Zarit label, the resulting supervision signal was almost certainly noisy given that not every turn produced by a caregiver with high overall burden expresses burden, and some turns from lower-burden caregivers plausibly do. This label noise likely explains the pattern observed, in which weak supervision achieved comparatively higher precision but lower recall than the baseline and LLM prompting, rather than uniformly outperforming them.

Third, the fully supervised model, despite being trained on substantially more data (191,508 human-coded sentences) and using a genuinely fine-grained (sentence-level) label, achieved the lowest F1-score. Only 1.1% of sentences were coded as expressing burden, reflecting a very high-class imbalance that is characteristic of rare-event detection tasks in naturalistic clinical dialogue. Because burden-relevant statements are inherently infrequent relative to the surrounding conversation, sentence-level classifiers may require imbalance-aware training strategies, richer contextual features (e.g., surrounding turns rather than isolated sentences), or a reformulation of the classification unit (e.g., grouping sentences into topical or temporal windows) to achieve acceptable recall without an unacceptable false-positive burden. As emphasized above, this result is not directly comparable to Methods 1 and 2, given the difference in label source and unit of analysis; it is presented to characterize the scale of the class-imbalance challenge rather than to rank Method 3 against Methods 1–2.

Several limitations should be considered. First, the parent study’s low initial inter-rater reliability (Cohen’s kappa of 0.25–0.30) indicates that early annotation may have been unreliable before protocol refinement; although agreement improved substantially for annotator pair #2 (kappa = 0.79), the generalizability of this improvement to all annotators and all transcripts should be confirmed. Second, the transcript-level held-out set used to evaluate Methods 1 and 2 was small (n = 30), limiting the precision of the reported performance estimates and the power to detect meaningful differences between methods. Third, the sample was drawn from a single implementation study (COPE in PACE) involving 17 dyads and 15 clinicians across 10 PACE sites in the United States; findings may not generalize to other care settings, caregiving populations, or dementia-care programs. Fourth, transcripts were produced by automatic speech recognition and may contain transcription errors, particularly around emotionally salient or disfluent speech, which could affect both human annotation and model performance. Fifth, using a single baseline (parent-study) Zarit score as the sole caregiver-level label for weak supervision does not capture within-caregiver variability in burden across sessions.

Taken together, these findings suggest that no single approach evaluated here is yet sufficiently accurate for unsupervised clinical deployment, but they point toward productive directions: combining LLM-based semantic reasoning with fine-grained, imbalance-aware supervised training; using active learning to prioritize annotation of likely burden-indicating segments; and validating burden predictions against measures beyond a single baseline Zarit score, such as repeated or session-level assessments.

## Conclusion

Detecting caregiver burden from transcribed clinical conversations remains a challenging NLP task^18^. Among the approaches evaluated, LLM prompting achieved the strongest transcript-level performance without requiring model fine-tuning, while a fully supervised sentence-level model was substantially limited by severe class imbalance. These results suggest that hybrid approaches that combine structured, prompt-based reasoning with fine-grained, imbalance-aware supervised learning and larger annotated corpora may represent a promising path toward clinically useful, ambient detection of caregiver burden. Continued investment in high-quality, reliably annotated datasets will be essential before such tools can responsibly support clinicians in identifying and addressing caregiver distress during routine care.

## Data Availability

Data produced in the present study may be available upon reasonable request to the authors.

## Acknowledgements

Research reported in this publication was supported by the National Institute on Aging of the National Institutes of Health under Award Number P30AG073105. The content is solely the responsibility of the authors and does not necessarily represent the official views of the National Institutes of Health. The authors acknowledge the important manual annotation work completed by Hannah Cho, PhD, Hanna Ramsberg, PhD, Freya Yang, PhD, and Emma Max, PhD.

